# Distinct metabolic features of genetic liability to type 2 diabetes and coronary artery disease: a reverse Mendelian randomization study

**DOI:** 10.1101/2022.04.13.22273833

**Authors:** Madeleine L Smith, Caroline J Bull, Michael V Holmes, George Davey Smith, Emma L Anderson, Joshua A Bell

**Author notes:** Corresponding author: Madeleine L Smith, Oakfield House, University of Bristol, UK, BS8 2BN.

## Abstract

Type 2 diabetes (T2D) and coronary artery disease (CAD) both have known genetic determinants, but the mechanisms through which their associated genetic variants lead to disease onset remain poorly understood. Here, we used large-scale metabolomics data to directly compare the metabolic features of genetic liability to T2D and to CAD. We performed two-sample reverse Mendelian randomization (MR) to estimate effects of genetic liability to T2D and CAD on 249 circulating metabolites from targeted nuclear magnetic resonance spectroscopy in the UK Biobank (N=118,466). We examined the potential for medication use to distort effect estimates by examining effects of disease liability on metformin and statin use and by conducting age-stratified metabolite analyses. Using inverse variance weighted (IVW) models, higher genetic liability to T2D was estimated to decrease high-density lipoprotein cholesterol (HDL-C) and low-density lipoprotein cholesterol (LDL-C) (e.g., HDL-C: -0.05 SD; 95% CI -0.07, -0.03, per doubling of liability), whilst increasing all triglyceride groups and branched chain amino acids (BCAAs). Estimates for CAD liability suggested an effect on reducing HDL-C as well as raising very-low density lipoprotein cholesterol (VLDL-C) and LDL-C, and LDL triglycerides. Liability to each disease was estimated to decrease apolipoprotein-A1, whilst only CAD liability was estimated with IVW to increase apolipoprotein-B (0.10 SD; 95% CI 0.03, 0.17). In pleiotropy-robust sensitivity models, T2D liability was still estimated to increase BCAAs, but several effect estimates for higher CAD liability reversed and supported decreased LDL-C and apolipoprotein-B. Estimated effects of CAD liability differed uniquely and substantially by age for non-HDL-C traits in particular, with, e.g., pleiotropy-robust models suggesting that higher CAD liability lowers LDL-C only at older ages when use of statins is common. Our results from pleiotropy-robust models support largely distinct metabolic features of genetic liability to T2D and to CAD, particularly higher BCAAs in T2D and lower LDL-C and apolipoprotein-B in CAD. Such apparently favourable effects of CAD liability differ substantially by age and likely reflect mediation by statin use in adulthood.

## INTRODUCTION

Globally, overall incidence rates of type 2 diabetes (T2D) and coronary artery disease (CAD) are increasing in parallel, together affecting over 500 million adults.(1,2) Both diseases have roots in disordered metabolism, but they differ in their life course development and clinical presentation. Genome-wide association studies (GWAS) have provided robust evidence that both T2D and CAD have genetic influences,(3,4) but the mechanisms through which the associated genetic variants increase disease risk remain poorly understood.

Recent advances in metabolomics technologies involve the simultaneous quantification of hundreds of molecular metabolites from a single biological sample, enabling detailed insight into the metabolic features of cardiometabolic diseases and their potential underlying mechanisms.(5) The application of metabolomics in observational epidemiological studies has helped to identify metabolites involved in the pathogenesis of T2D, or that might contribute to T2D incidence.(6–16) For example, a recent meta-analysis identified associations of incident T2D with differences in several lipid and non-lipid metabolites, including higher levels of branched chain amino acids (BCAAs), some fatty acids and acylcarnitines, amongst others.(17) For CAD, metabolomics in observational studies has helped to identify key metabolic characteristics of the disease,(18–25) to predict and diagnose CAD,(26–29) and to predict outcomes and mortality in CAD patients.(30–32) However, observational studies are limited in their ability to infer whether metabolic traits are causal, rather than just predictive for disease, as these studies are liable to bias due to residual confounding by lifestyle factors and other diseases.

Mendelian randomization (MR), a genetic epidemiological approach that uses genetic variants associated with modifiable exposures to estimate the unbiased effects of such exposures on outcomes,(33) has been applied to investigate associations between lipid levels and T2D and CAD (34–37), with the two diseases appearing to have differing lipid aetiology: higher low-density lipoprotein cholesterol (LDL-C) and triglycerides increase CAD risk, whereas higher LDL and high-density lipoprotein cholesterol (HDL-C) reduce T2D risk.(36) More recently, MR has been used in ‘reverse’ to estimate effects of genetic liability for disease and potential biomarkers, to reveal features of the developing disease process, which include both causal and non-causal factors for disease incidence.(38) Using reverse-MR, higher genetic liability to T2D diagnosed in adulthood has been estimated to decrease total lipid content in HDL particles, as well as increase BCAA and inflammatory glycoprotein acetyl levels, as early as 8 years of age.(39) Higher genetic liability to CAD has also been estimated to affect circulating metabolite levels in early life, especially higher very-low-density lipoprotein (VLDL) and total lipids in LDL.(40) These MR studies in children have the advantage of low disease prevalence (and therefore little reverse causality), and low medication use, so are useful for identifying causal factors. Studies in older adults are better for identifying predictors of disease, as there is a greater burden of prevalent disease, however increased medication use with age has the potential to distort results. Previous reverse-MR studies of the impact of genetic liability to T2D or CAD on metabolic traits are limited by their small sample sizes; cohort studies of ~5,000 participants or GWAS of less than ~25,000 adults. Moreover, the metabolic profiles of genetic liability to T2D and CAD have not been directly compared within the same analytical setting, and so the ways in which these profiles differ have not yet been revealed.

Here, we compare the metabolic features of genetic liability to T2D and to CAD using a reverse-MR framework, with the aim of revealing the pattern of metabolic traits which characterise the development of each disease. We used the largest, most recent summary-level GWAS data on circulating metabolites measured using targeted metabolomics (N~120,000) from the population-based UK Biobank cohort study, representing a 5-fold larger sample size than previous studies. To examine the potential for effect estimates to be distorted by medication use, we examined the effects of T2D and CAD liability on the use of metformin and statins, and repeated analyses using data from age stratified GWAS of metabolites, given that medication use differs markedly by age. We also estimated the effects of disease liability on more distal lifestyle-related factors, including adiposity, smoking, and alcohol consumption.

## METHODS

### Genetic instruments for T2D liability and CAD liability

#### T2D liability

We identified single nucleotide polymorphisms (SNPs) that were independently associated with T2D at P < 5 × 10^−8^ from a large-scale GWAS meta-analysis combining data from 32 GWAS, including 74,124 T2D cases and 824,006 controls of European ancestry.(3) Across included studies, female sex ranged from 37 to 64% and mean (SD) age range was 23.9 (2.1) to 61.3 (2.9) years. T2D was diagnosed across included studies using a range of criteria, including use of oral diabetes medication, diagnostic fasting glucose or HbA1c levels, or self-report. The T2D GWAS was conducted with and without adjustment for BMI, and we used the 231 SNPs identified in the non-BMI-adjusted GWAS to reduce potential for collider bias.(41) For variants that were in linkage disequilibrium (LD) based on R^2^ > 0.001, the SNP with the lowest P-value in each set of variants in LD was retained. This left 167 independent SNPs associated with T2D (**Supplementary Table 1**) for inclusion in our reverse-MR analysis.

#### CAD liability

SNPs for CAD were identified in a large-scale GWAS meta-analysis of 10 GWAS conducted among middle-aged adults, with study-specific sample and variant filters applied and adjustment for study-specific covariates. The study had a total sample of 181,522 cases among 1,165,690 study participants, who were 46% female and largely of European ancestry (>95%). Potential ancestry effects were accounted for by transethnic comparison with a GWAS from Biobank Japan. Case status was defined based on relevant hospital codes for prevalent or incident CAD. The GWAS detected 241 genetic variants independently associated with CAD at P < 5 × 10^−8^. For variants that were in LD based on R^2^ > 0.001, the SNP with the lowest P-value in each set of variants in LD was retained, leaving 145 SNPs (**Supplementary Table 1**).

### Outcome data on circulating metabolites and medication use

Summary statistics from GWAS of circulating metabolites conducted among European participants of the UK Biobank study were used. Details of the UK Biobank design, participants, genomic quality control (QC) and its strengths and limitations have been reported previously.(42–44) Briefly, 502,549 adults aged 40-69 years were recruited between 2006-2010 via 22 assessment centres across England, Wales, and Scotland. Non-fasting EDTA plasma samples from a random subset of participants (N = 118,466) were analysed for levels of 249 metabolic traits (165 concentrations and 84 derived ratios) using targeted high-throughput proton nuclear magnetic resonance (^1^H-NMR) spectroscopy (Nightingale Health Ltd; biomarker quantification version 2020).(45) These metabolic traits comprised routine lipids, lipoprotein subclass profiling, fatty acid composition and various low-molecular-weight metabolites including BCAAs, ketone bodies, glycolysis-related traits and inflammatory glycoprotein acetyls. All metabolic trait measurements were normalized and standardised prior to analyses using rank-based inverse normal transformation. Metabolite GWAS were adjusted for genotype array, sex, and fasting time. We also included C-reactive protein (CRP) for comparison with glycoprotein acetyls, measured in UK Biobank via the same blood samples, analyzed by immunoturbidimetric-high sensitivity analysis on a Beckman Coulter AU5800 (N = 469,772).

Statins and metformin are commonly prescribed medications for the prevention/treatment of CAD and T2D, respectively, particularly among adults identified as having higher risk.(46) These medications can alter the levels of circulating metabolites (47,48) and thus may distort associations of genetic liability to T2D and CAD with metabolites (via mediation). We therefore additionally estimated the effects of T2D and CAD liability on the self-reported use of statins (atorvastatin and simvastatin) and metformin obtained from a sample of 462,933 UK Biobank participants using interviews, self-reporting and medical records (UK Biobank Data-Field 20003), to inform the interpretation of metabolite results. These medication outcomes also served as positive controls, with liability to CAD expected to most strongly raise statin use, and liability to T2D expected to most strongly raise metformin use.

For the purposes of further examining distortions by medication use, we used metabolite GWAS on the same sample which were divided into age tertiles (containing ages: 39-53 years, 53-61 years, and 61-73 years), and which used the same standardisations and covariate adjustments. Based on previous work in UK Biobank,(49) the estimated prevalence of statin medication use within these same tertiles (youngest to oldest) is 5%, 17%, and 29%, respectively.

Data used for additional outcomes (adiposity, smoking and alcohol consumption) is described in **Supplementary File**.

### Statistical approach

To generate MR estimates of the effect of genetic liability to T2D and CAD on metabolites (with ages combined and within age tertiles), and on medication use and additional outcomes noted above, we integrated estimates of the association of SNPs with exposures (T2D and CAD separately), with estimates of the association of those same SNPs with each metabolic trait measured in UK Biobank. Summary statistics were harmonised using the ‘harmonise_data’ function within the TwoSampleMR package.(50) Using the same package, we used inverse variance weighted (IVW) regression in the main analysis, which assumes that none of the SNPs are pleiotropic (exclusion restriction), as well as the two other core IV assumptions: relevance and independence.(51) To examine and correct for potential horizontal pleiotropy, we used 3 additional sensitivity methods for each exposure-outcome pair: MR-Egger (which allows for all SNPs to be pleiotropic),(52) weighted median (which allows for up to half of the weighted SNPs to be pleiotropic and is less influenced by outliers than other models)(51), and the weighted mode (which assumes that the most common effect is consistent with the true causal effect, i.e. not biased by horizontal pleiotropy).(53) To aid interpretation, all estimates and standard errors were multiplied by 0.693 (log_e_2) and represent the normalised SD unit difference in each outcome trait (metabolites, medications, adiposity, smoking, and alcohol) per doubling of genetic liability to T2D or CAD, based on previous recommendations(54). Statistical analyses were performed in R version 4.0.2.

## RESULTS

Instrument count and strength are reported in **Supplementary File**.

### Lipids and lipoproteins

IVW estimates for the effect of higher liability to T2D and to CAD on lipids tended to differ between diseases (**Figure 1**). Higher T2D liability was estimated to decrease total, HDL-C and LDL-C (e.g., HDL-C -0.05 SD; 95% CI -0.07, -0.03), with no strong evidence of an effect on VLDL-C. IVW estimates for CAD liability on cholesterol were less precise but suggested an effect on lowering HDL-C and raising total-C, VLDL-C and LDL-C. T2D liability appeared to increase all triglyceride groups (e.g., triglycerides in VLDL 0.04 SD; 95% CI 0.01, 0.07), whilst CAD liability had little effect on triglyceride groups except for LDL triglycerides which were increased. T2D liability was estimated to increase VLDL particle size (0.04 SD; 95% CI 0.02, 0.07), and decrease LDL and HDL particle size (−0.04 SD; 95% CI -0.06, -0.02 for both), whilst CAD liability was estimated to increase LDL particle size (0.06 SD; 95% CI 0.02, 0.10), and decrease HDL particle size (−0.06 SD; 95% CI -0.11, -0.01). T2D liability appeared to decrease, whilst CAD liability appeared to increase, apolipoprotein B, although estimates were imprecise for CAD; whilst liability to either disease appeared to decrease apolipoprotein A1 (T2D -0.03 SD; 95% CI -0.05, -0.01; CAD -0.07 SD; 95% CI -0.11, -0.03). Effect estimates across sensitivity models were imprecise for T2D and CAD with each lipid trait (e.g., MR Egger estimate for effect liability to T2D on total-C 0.00 SD; 95% CI -0.05, 0.04; Egger P-values largely >0.05). Of note, the effect of higher CAD liability appeared inverse for VLDL-C, LDL-C, and apolipoprotein B in weighted median and weighted mode models (**Supplementary Tables 2-4; Supplementary Figure 6**). Evidence of heterogeneity was strong for all lipid and lipoprotein traits, e.g., Cochran’s Q P-value = 4.22e^-188^ for the IVW estimate for T2D liability on total-C (**Supplementary Table 2**). Where the direction of effect estimates was inconsistent across MR models, we examined scatter plots of individual SNP effects (**Supplementary Figure 4**). These supported the existence of several outlier SNPs which were biasing estimates from IVW and MR Egger models; estimates from weighted median and weighted mode models appeared not to be influenced by these outliers and were in line with the majority of SNP effects.

**Figure 1:**
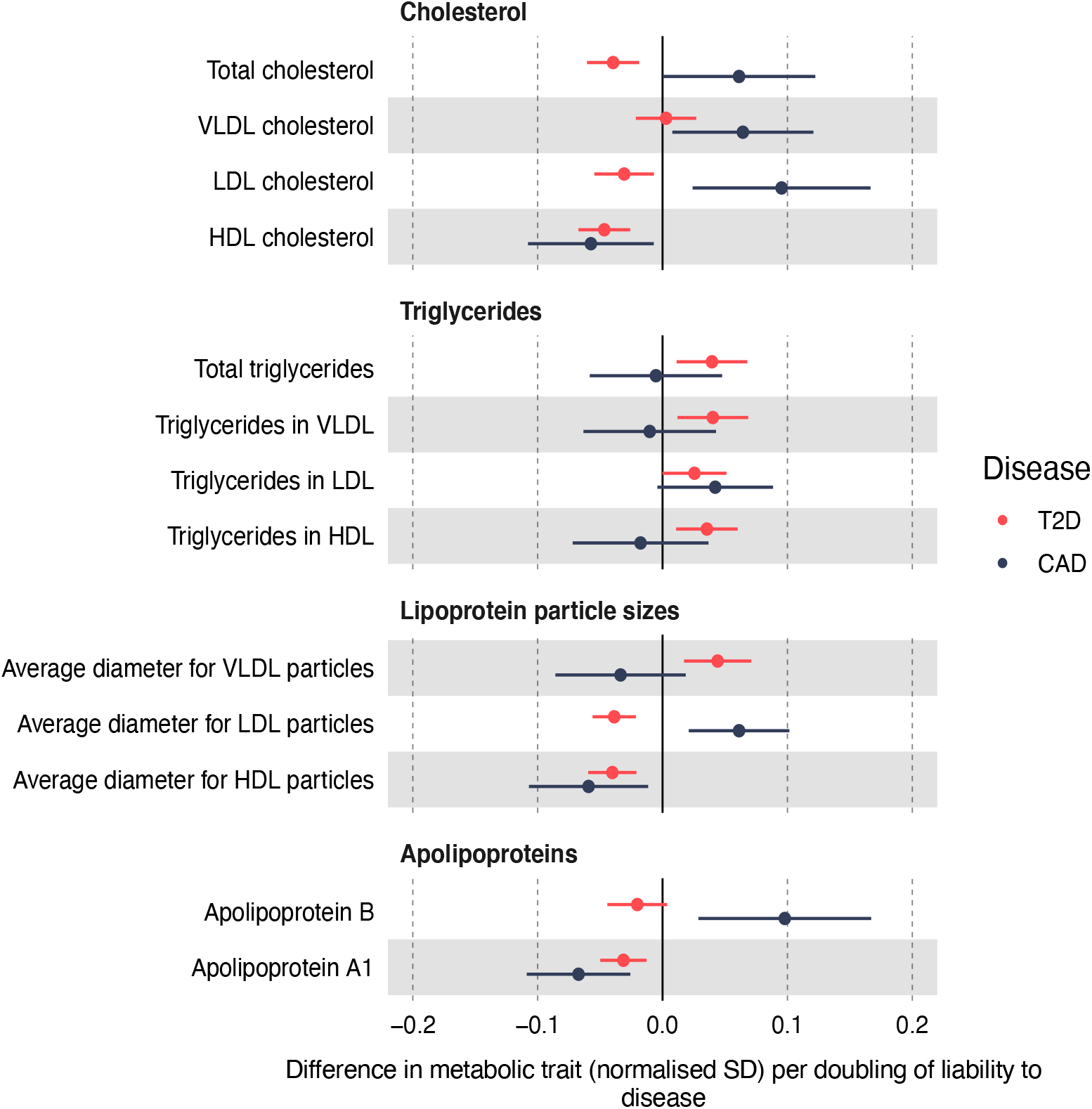
Effect of T2D and CAD liability on lipids and lipoproteins. Effect estimates are normalised SD unit differences in metabolite per doubling of liability to T2D or CAD based on IVW models.

The magnitudes of the effect of liability to T2D or CAD on statin or metformin use were small. As expected, higher T2D liability most strongly increased odds of taking metformin (**Supplementary Figure 1**; OR 1.02; 95% CI 1.01, 1.02), whilst higher CAD liability most strongly increased odds of taking simvastatin (OR 1.03; 95% CI 1.03, 1.04). The effect of higher T2D liability on metformin use was robust across weighted median and weighted mode models but attenuated in the MR-Egger model (Egger intercept P-value=6.09e^-16^ (**Supplementary Tables 3&4; Supplementary Figure 5**)). Estimates of CAD liability for atorvastatin and simvastatin use were consistent across sensitivity models (**Supplementary Table 2**).

The estimated effects of T2D and CAD liability on several lipids, particularly non-HDL cholesterol, differed markedly by age (**Figure 2**). Among these age-stratified results, some estimated effects of T2D liability on lipids were inconsistent across sensitivity models, including for apolipoprotein B and LDL-C (**Supplementary Figure 7**), however, there was little evidence that Egger intercepts for these estimates differed from zero (**Supplementary Table 6**). Based on outlier-robust weighted median and mode models, higher CAD liability was estimated to decrease LDL-C and apolipoprotein B within the oldest age tertile (statin use 29%); these effects diminished within the intermediate age tertile (statin use 17%) and were null within the youngest age tertile (statin use 5%).

**Figure 2.**
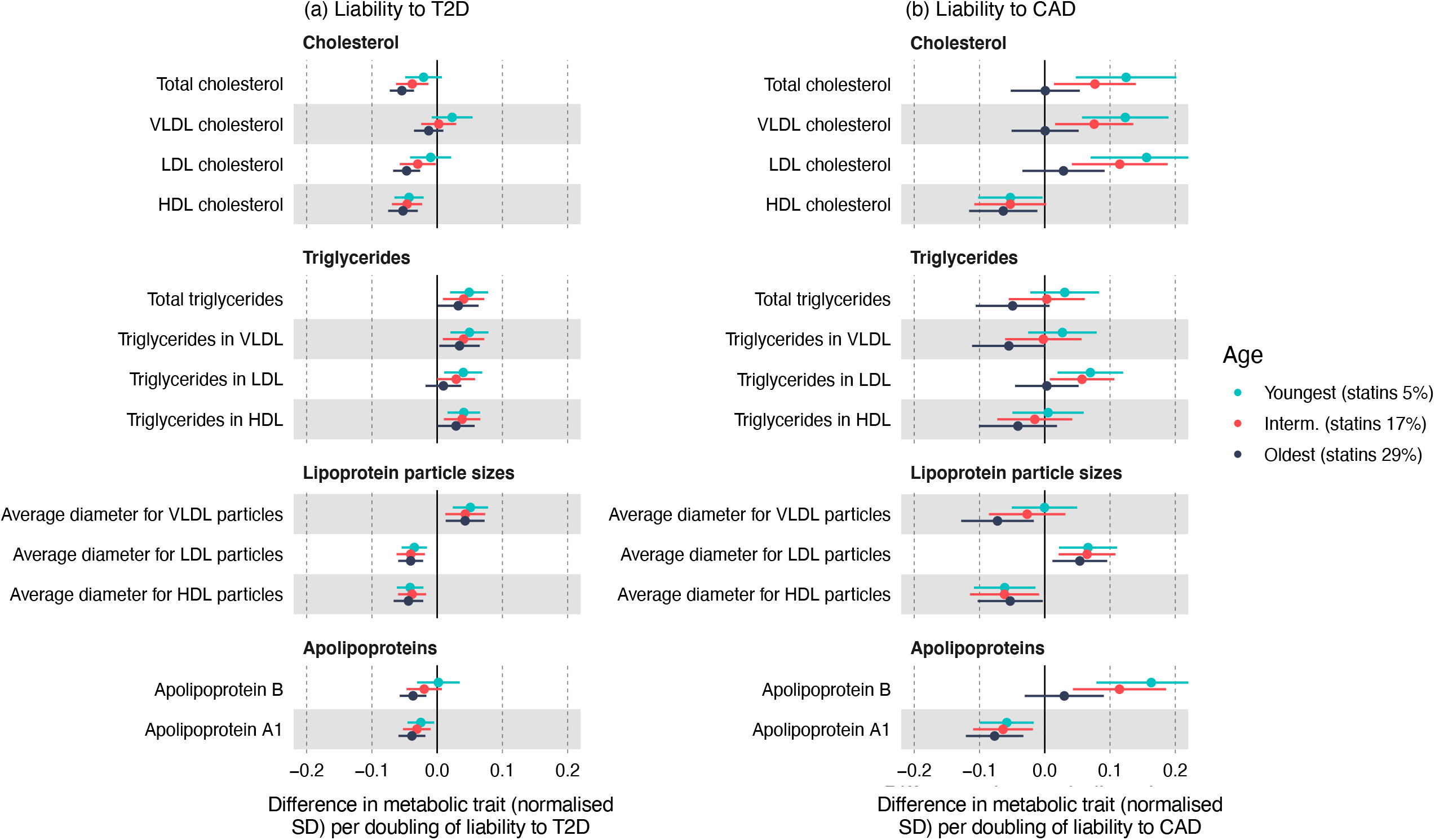
Effect of T2D (a) and CAD (a) liability on lipids and lipoproteins in age tertiles. Effect estimates are normalised SD unit differences in metabolite per doubling of liability to disease based on IVW models.

### Fatty acids and amino acids

Effects on fatty acid traits were largely opposite for T2D liability and CAD liability (**Figure 3**). For example, higher T2D liability was estimated to decrease the ratio of docosahexaenoic acid to total fatty acids (−0.02 SD; 95% CI -0.03, -0.01) whereas CAD liability was estimated to increase the same ratio (0.03 SD; 95% CI 0.00, 0.05). CAD liability was estimated to decrease the ratio of saturated to total fatty acids, and increase the ratio of docosahexaenoic acid to total fatty acids, and both of these were consistent across sensitivity models. Higher T2D liability was estimated to increase all amino acids (except for glycine), including total BCAAs (IVW 0.05 SD; 95% CI 0.04, 0.07), which was robustly positive across sensitivity models (**Supplementary Figure 5**). There was consistently little evidence of an effect of CAD liability on any amino acid, except for total BCAAs and valine for which MR-Egger estimates suggested a decrease (**Supplementary Tables 2-4; Supplementary Figure 7**). Evidence of heterogeneity was strong for most fatty acid and amino acid traits (**Supplementary Table 2**).

**Figure 3:**
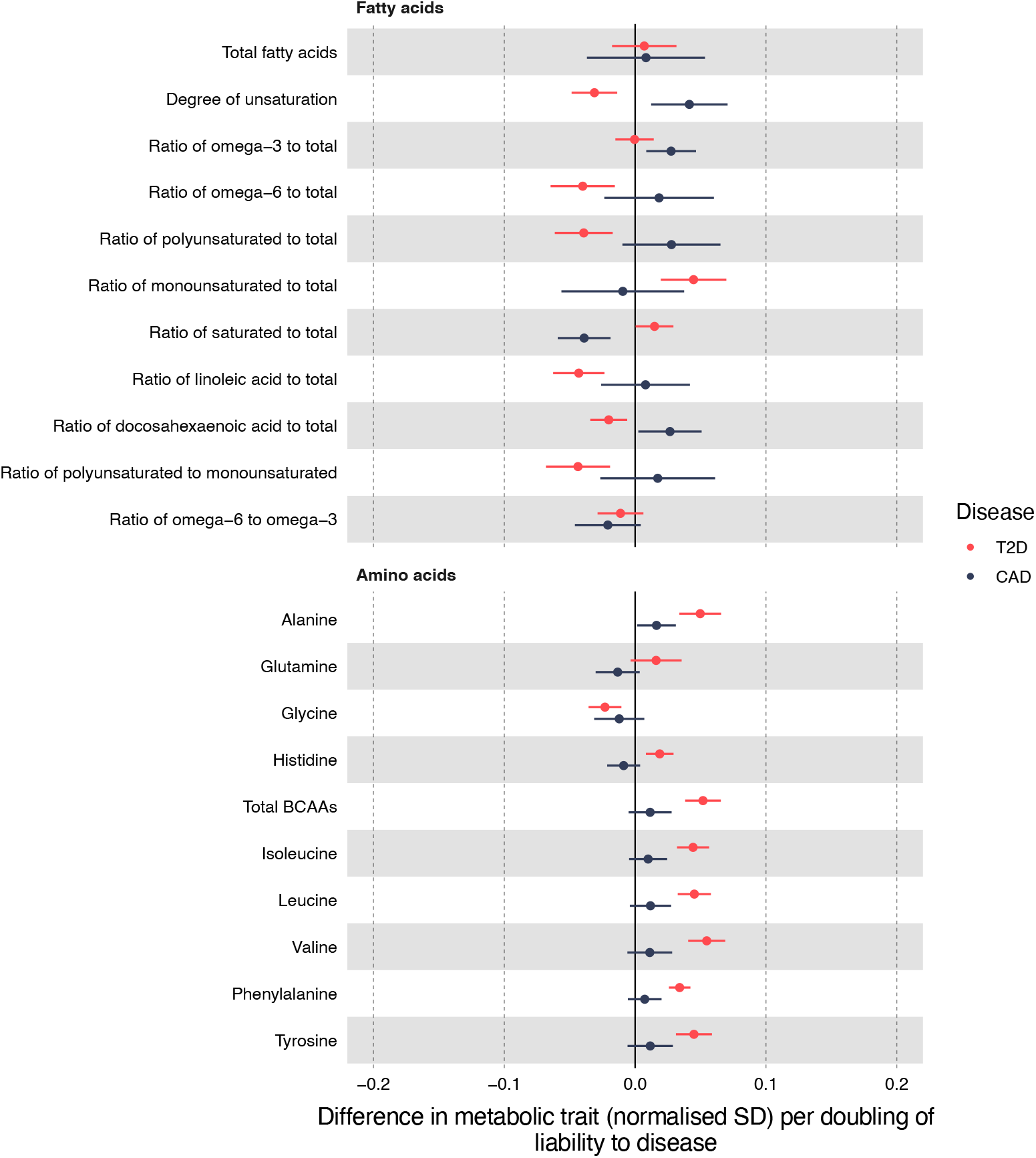
Effect of T2D and CAD liability on fatty acids and amino acids. Effect estimates are normalised SD unit differences in metabolite per doubling of liability to T2D or CAD based on IVW models.

The effects of T2D liability on fatty acids and amino acids as estimated by IVW models were fairly consistent with age, exceptions including total fatty acids, ratio of saturated to total fatty acids, and histidine (**Figure 4**). For histidine, estimates were consistently null in the intermediate tertile, and consistently positive in the oldest and youngest tertiles (**Supplementary Tables 5-7**). The effect of higher CAD liability on total fatty acids decreased with age, with the youngest tertile exhibiting an increase, the intermediate tertile a null effect, and the oldest tertile a decrease (**Supplementary Table 5**). The effect of higher CAD liability on both omega-3 to total fatty acids and docosahexaenoic acid to total fatty acids ratios was null in the youngest tertile but positive in both the intermediate and oldest tertiles (consistent across sensitivity models (**Supplementary Tables 5-7**)).

**Figure 4.**
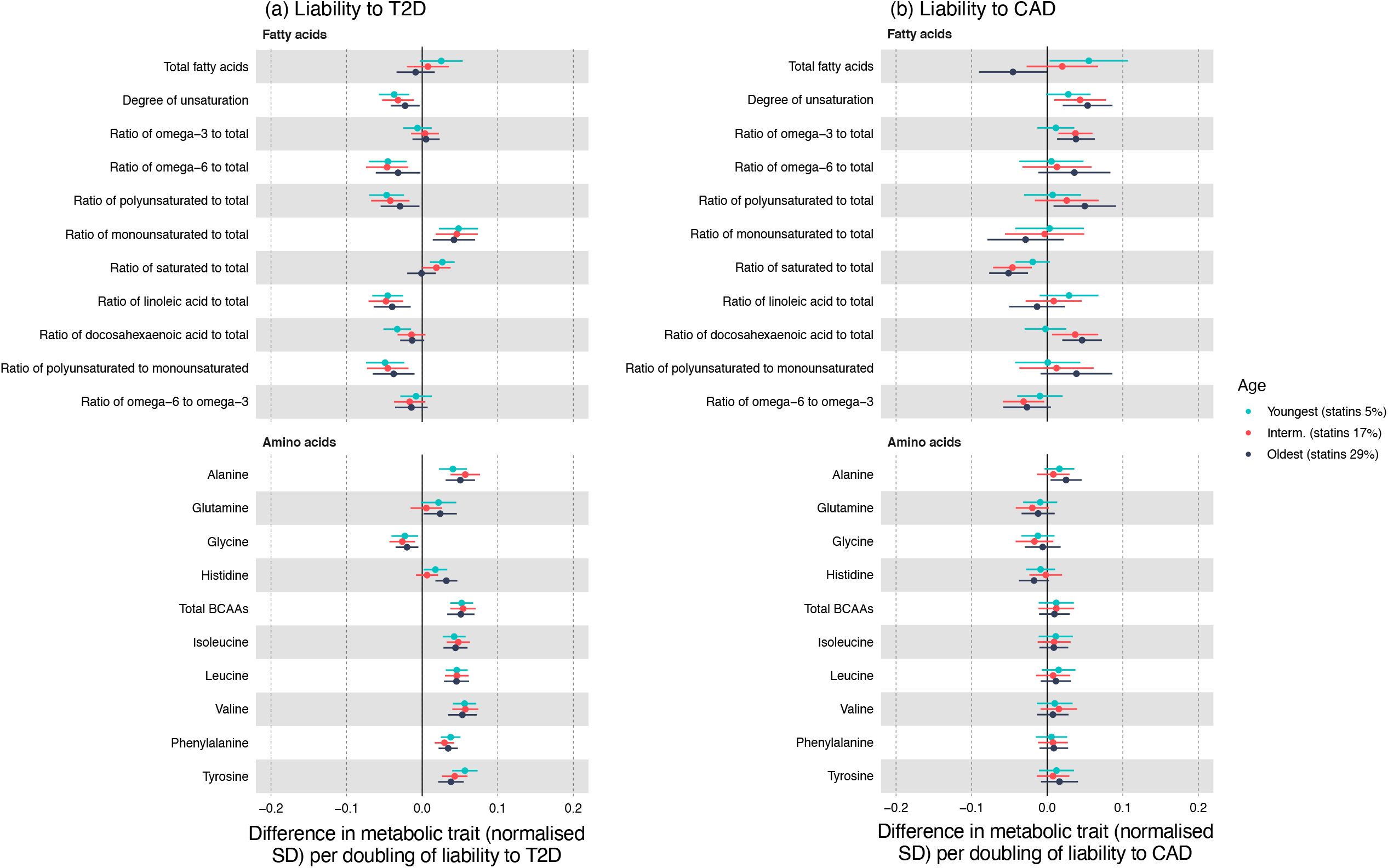
Effect of T2D (a) and CAD (b) liability on fatty acids and amino acids in age tertiles. Effect estimates are normalised SD unit differences in metabolite per doubling of liability to disease based on IVW models.

### Glycolysis traits and other metabolites

As expected, T2D liability consistently increased glucose (0.08 SD; 95% CI 0.06, 0.10) across sensitivity models (**Supplementary Tables 2-4; Supplementary Figure 5**); yet there was no effect of CAD liability on glucose. There was little evidence of effect of T2D liability or CAD liability on other glycolysis related metabolites, ketone bodies or fluid balance metabolites (**Figure 5**). Only T2D liability was estimated to slightly increase glycoprotein acetyls (IVW 0.02 SD; 95% CI 0.00, 0.04), although there were inconsistencies across models (**Supplementary Figure 5**). Effect estimates for CRP were similar to those for glycoprotein acetyls. Evidence of heterogeneity was strong for most pre-glycaemic and other metabolic traits (**Supplementary Table 2**).

**Figure 5:**
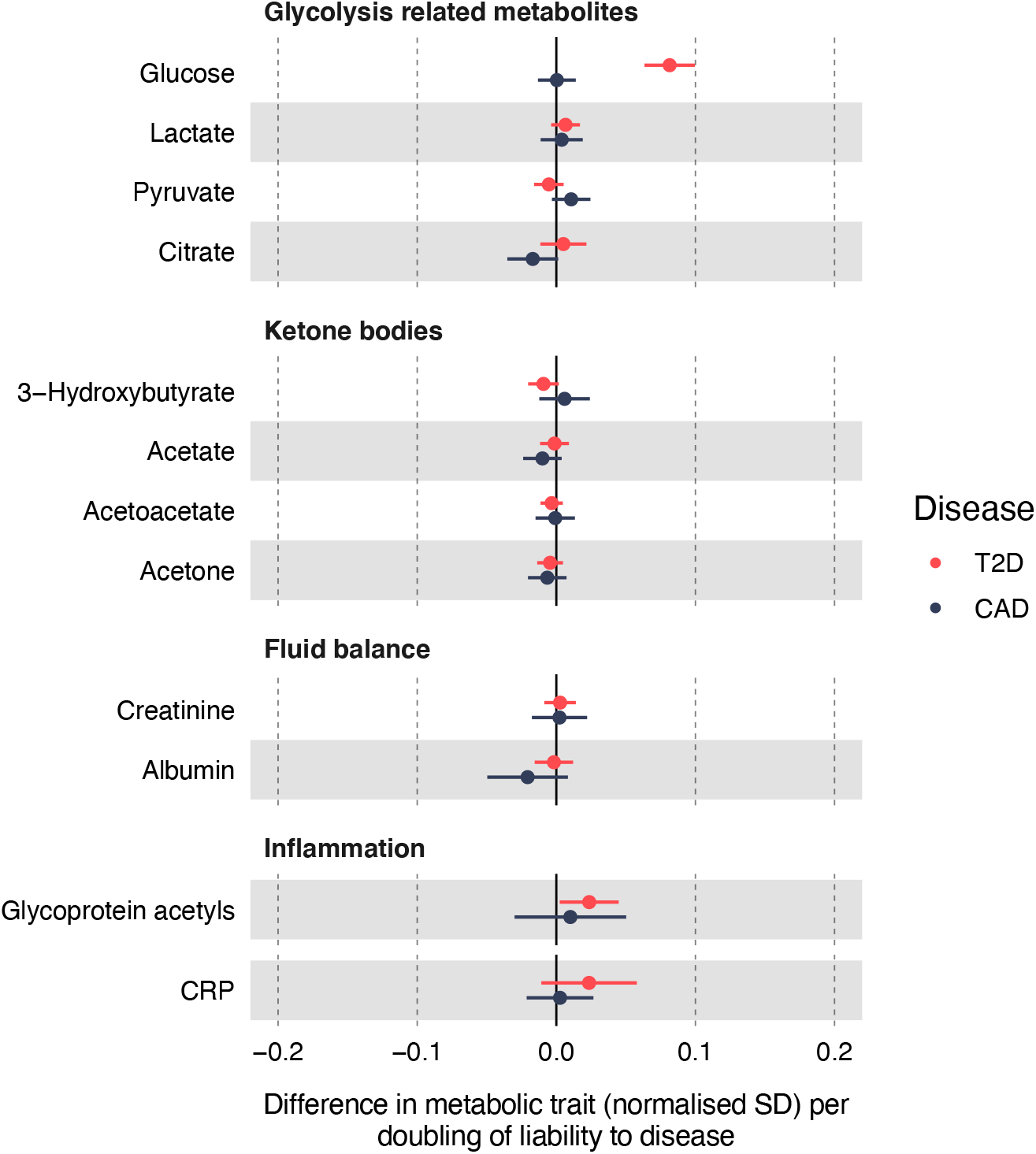
Effect of T2D and CAD liability on glycolysis related metabolites, ketone bodies, fluid balance metabolites and inflammation metabolites. Effect estimates are normalised SD unit differences in metabolite per doubling of liability to T2D or CAD based on IVW models.

Estimates of the effects of T2D and CAD liability were largely consistent across age tertiles for glycolysis traits and ketone bodies, except for CAD liability on lactate and pyruvate which was positive in the youngest and null in the intermediate and oldest tertiles (**Figure 6**). The effect of higher T2D liability on glycoprotein acetyls was positive in the youngest (IVW 0.04 SD, 95% CI 0.02, 0.06) and null in the oldest tertile. This was not robust across sensitivity models (**Supplementary Tables 6-7; Supplementary Figure 9**).

**Figure 6.**
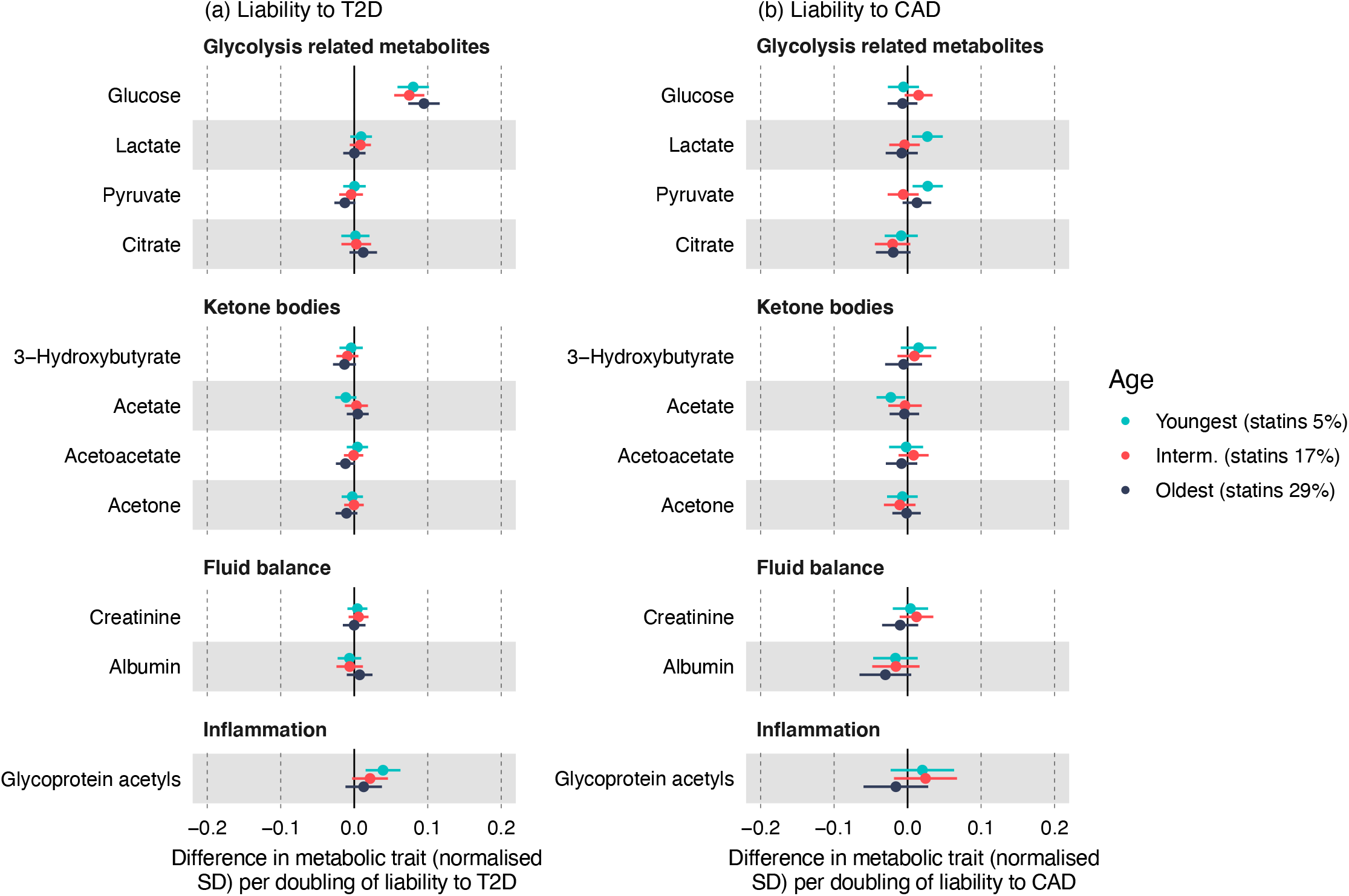
Effect of T2D (a) and CAD (b) liability on glycolysis related metabolites, ketone bodies, fluid balance metabolites and glycoprotein acetyls in age tertiles. Effect estimates are normalised SD unit differences in metabolite per doubling of liability to disease based on IVW models.

### Adiposity, smoking, and alcohol consumption

Higher genetic liability to T2D was estimated to increase all adiposity outcomes according to IVW estimates (**Supplementary Figure 2**; e.g., BMI: 0.03 SD; 95% CI 0.01, 0.05). However, there was evidence to suggest those estimates may be biased by horizontal pleiotropy; MR-Egger P-values were <0.05 for all adiposity traits (**Supplementary Table 3)**. There was little evidence of an effect of liability to CAD on any of the adiposity outcomes according to IVW, weighted median and weighted mode estimates, however MR-Egger estimates suggested a small effect on decreased adiposity (e.g., fat mass: -0.03 SD; 95% CI -0.07, 0.00). Evidence of heterogeneity was strong for all adiposity traits (**Supplementary Table 2**). The effects of T2D and CAD liability on both smoking status and alcohol drinking status were null (**Supplementary Figure 3)**. Higher T2D liability was estimated to increase alcohol intake frequency in IVW models (0.02 SD, 95% CI 0.01, 0.04); this was null in sensitivity models (**Supplementary Tables 3&4**). Higher CAD liability had no effect on alcohol intake frequency. There was no effect of liability to either disease on pack years of smoking.

### Summary XY plot of T2D and CAD liability for metabolic and other outcomes

Estimates of the effects of T2D and CAD liability on each metabolite (ages combined), adiposity, and medication use were visualized in an XY plot to compare the overall pattern of effects of liability to either disease across outcomes (**Figure 7**). To better visualize the comparison across different feature classes, only a subset of metabolites is included on the XY plot (the same as included across **Figures 1-6**); the slope of this regression line was -0.16 with an R^2^ of 0.02. When including all 266 features (all metabolites, adiposity, smoking, alcohol, CRP), the slope of the regression line was -0.49 with an R^2^ of 0.11. Together, this indicates a weak association between profiles, giving further evidence that genetic liability to T2D and to CAD have distinct metabolic features.

**Figure 7.**
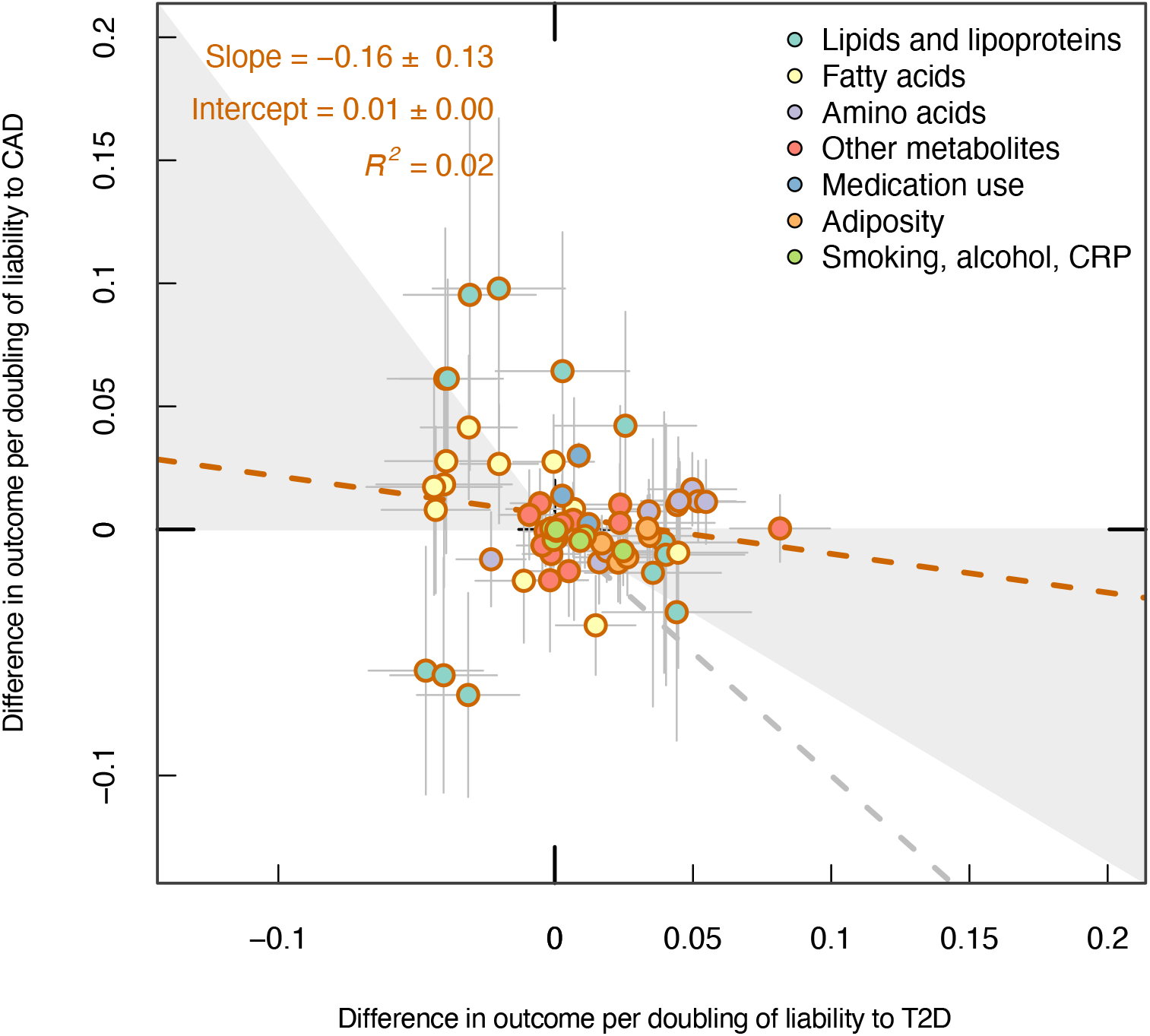
Comparison of the effects of T2D and CAD liability on adiposity, metabolic traits, medication use, smoking and alcohol use. Effect estimates are SD unit differences in adiposity or metabolic trait, or logodds for medication use and smoking/alcohol status, per doubling of odds of T2D or CAD, based on IVW models.

## DISCUSSION

In this study, we directly compared the metabolic profiles of genetic liability to T2D and CAD, two diseases which commonly co-occur but which involve different pathophysiology and clinical presentation. We applied a reverse-MR framework(38) using new summary-level GWAS data on metabolomics from UK Biobank, which enabled a 5-times larger sample size over previous studies.(39,40) Our results based on pleiotropy-robust models suggest that genetic liability to T2D and to CAD have largely distinct metabolic features, including increased BCAAs in T2D and decreased LDL-C and apolipoprotein B in CAD. Such apparently favourable effects of CAD liability differ substantially by age and likely reflect mediation by statin use in adulthood.

Statins and metformin are commonly used for the prevention/treatment of CAD and T2D, respectively. The overall prevalence of statin use in UK Biobank is 16%, with men almost twice as likely to be taking statins as women, and use increasing markedly across age tertiles.(49) Statins and metformin are known to lower LDL-C (46,55) and will likely have direct and indirect effects on other metabolic traits.(47) As expected, our results suggest that increased T2D liability most strongly increases the odds of metformin use whilst increased CAD liability most strongly increases the odds of statin use, providing positive controls and suggesting specificity of genetic instruments used for each disease. Medication use may therefore modify the effect of T2D or CAD liability on metabolic traits and distort results in the form of underestimated, or even reversed, effects of genetic liability. Our results based on (non-pleiotropy-robust) IVW models suggest that higher CAD liability increases LDL-C and apolipoprotein B which is consistent with a previous reverse-MR conducted across younger samples,(40) suggesting that perturbations in LDL-C in early life due to CAD liability persist into adulthood. We did, however, see attenuation of these effects in the oldest age tertile which could be due to the increased statin use in the older sample; and the direction of effect was inconsistent across pleiotropy-robust sensitivity models, suggesting that these particular results were vulnerable to outlier SNP effects. Results based on pleiotropy/outlier-robust weighted median and mode models suggested that CAD liability has an inverse effect on LDL-C, VLDL-C, and apolipoprotein B; this again differed substantially by age, with inverse effects only at older ages, and with attenuated or null effects at younger ages. Medication use may explain inconsistencies between results for T2D or CAD liability and LDL-C from this study and previous studies, particularly studies of younger people who are less likely to be taking medication.(40,56) By stratifying the cohort by age as a proxy for medication use before performing metabolite GWAS, we were likely better positioned to overcome these distortions and isolate the effects of disease liability. Conditioning on age (as a proxy) instead of medication use itself, which is affected by disease liability, also reduced the potential for collider bias.

Increasing HDL-C has been hypothesized as a mechanism to reduce CAD risk, based largely on conventional non-genetic epidemiology studies.(57) However, evidence from MR studies has shown that HDL-C-raising genetic variants do not reduce CAD risk,(37,58) and a meta-analysis showed that that HDL-C modifying treatments did not reduce cardiovascular mortality.(59) Our current study detected an effect of CAD liability on decreasing HDL-C levels, suggesting that reduced HDL-C is an early (likely non-causal) feature of CAD development, potentially explaining why lower HDL-C appears to be associated with higher CAD risk in observational settings. We also found some evidence to suggest that both LDL-C and HDL-C are reduced in response to increased T2D liability, which is consistent with evidence from a previous reverse-MR study among young people for HDL-C but not LDL-C (which was raised),(39) and a conventional ‘forward’-MR study that found increased HDL-C and LDL-C decreased risk of T2D.(36) This is also in accordance with evidence that statin therapy (which lowers LDL-C) increases risk of T2D.(60) However, there were inconsistencies in estimates for effect on these traits across pleiotropy-robust sensitivity models.

Our results suggest that T2D and CAD liability confer opposite effects on fatty acid metabolism, which were largely consistent across pleiotropy/outlier-robust sensitivity models. Inverse effects of T2D liability on various fatty acid metabolites (e.g., omega-6 and polyunsaturated) support them as potential candidates for further functional studies into their role in protection against T2D. However, a systematic review of randomized controlled trials of polyunsaturated fatty acid supplementation found no strong evidence that increasing omega-3, omega-6 or total polyunsaturated fatty acids provided protection against or treated T2D.(61) We observed an effect of higher CAD liability on increasing ratios of omega-3 fatty acids and docosahexaenoic acids to total fatty acids. This contrasts with existing evidence of a protective effect of omega-3 fatty acids on cardiovascular outcomes.(62) We saw evidence of substantial age differences in these effects, where higher CAD liability increased these ratios in the older tertiles but not the youngest, which could be an effect of medication or improved lifestyle in response to a CAD diagnosis.

Consistent with previous studies, we found consistent evidence across ages and sensitivity models that T2D liability robustly increased total BCAAs(17,39) whereas there was no evidence of an effect of CAD liability on BCAAs, highlighting that increased total BCAAs are exclusively a feature of T2D liability. Although both T2D and CAD are associated with inflammation,(63) evidence of an effect on levels of glycoprotein acetyls from this study was not robust for liability to either disease when viewing ages collectively; however, higher T2D liability did appear to modestly raise glycoprotein acetyls within the youngest age group. It is difficult to draw conclusions about the role of inflammation in either disease process from our study, however, given that we only considered two inflammatory markers, omitting other potentially important ones such as interleukin-6 and interleukin-1β that may play a role in disease pathogenesis.(64,65)

It is well established that excess adiposity is a causal risk factor for T2D,(66) and there is strong causal evidence that adiposity increases CAD risk in conventional (forward-direction) MR studies(67). Presently, we did not find consistent evidence that either T2D or CAD liability raises adiposity. This likely reflects the more distal nature of adiposity as compared with circulating metabolites, and possibly highly pleiotropic variants among the outcome SNPs.(68) We also did not find consistent evidence that either T2D or CAD liability increases smoking or alcohol behaviours. Although increased smoking and drinking are risk factors for both T2D and CAD,(69,70) the lack of signal seen in our study suggests that they are more distal features of disease liability. This provides further evidence that the reverse-MR approach is useful for revealing more proximal factors such as metabolites that are directly involved in the disease process and highlights the importance of combining evidence from MR study designs in different directions to reveal the full scope of factors contributing to disease development.

### Study limitations

The samples used to generate genetic instruments for T2D and CAD included participants from UK Biobank, the same sample from which we obtained genetic association estimates for metabolic traits, medication use, and lifestyle-related outcomes (~26% and ~48% overlap, respectively). This may have led to bias in the results in the direction of the observational association, however, given the strong genetic instruments for T2D and CAD liability, bias from sample overlap is likely small.(71) Another limitation is the unrepresentative nature of UK Biobank (initial response rate ~5%) and therefore is vulnerable to various forms of selection bias. Replication of this study in other large cohort studies and application of different approaches will allow more robust metabolic characterisation of T2D and CAD liability, although UK Biobank is currently the largest such data in existence. The metabolite GWAS with all ages collectively was not age-adjusted, leaving the potential for distortions by age within those metabolite effect estimates; although we were able to additionally examine these effects within age tertiles. Use of only summary-level data limits the capacity to fully explore effects of other factors such as medication use, sex, and ethnicity, which may influence metabolites. A further limitation is the reliance on targeted NMR metabolomics, rather than mass spectrometry which is not yet available at large scale but offers a broader representation of metabolites beyond lipid subclasses; targeted NMR is often considered more clinically relevant, however. The smoking pack-years GWAS was restricted to ever-smokers, which may induce sampling bias and invalidate the MR assumptions for that analysis.

### Conclusions

Our results support largely distinct metabolic profiles of genetic liability to T2D and to CAD using metabolite GWAS data from the largest cohort to date. Our most robust findings, based on pleiotropy-robust sensitivity models, suggest that T2D liability increases BCAAs, whilst CAD liability decreases LDL-C and apolipoprotein B. Such apparently favourable effects of CAD liability differ substantially by age and likely reflect mediation by statin use in adulthood.

## Data Availability

All data produced in the present study are available upon reasonable request to the authors

## ACKNOWLEDGEMENTS

We thank the participants of the UK Biobank study and the genome-wide association study consortiums who made their summary statistics publicly available for this study.

## DATA AVAILABILITY

All summary level GWAS results are publicly available through the IEU-OpenGWAS platform, accessible at https://gwas.mrcieu.ac.uk/.

## AUTHOR CONTRIBUTIONS

JAB, ELA and CJB conceived and planned the study and supervised analyses. MLS conducted the analyses and wrote the first draft. All others critically reviewed the intellectual content of manuscript drafts and with MLS approved the final version for submission.

