## Supplementary Figures 1-12 for "Distinct metabolic features of genetic liability to type 2 diabetes and coronary artery disease: a reverse Mendelian randomization study"

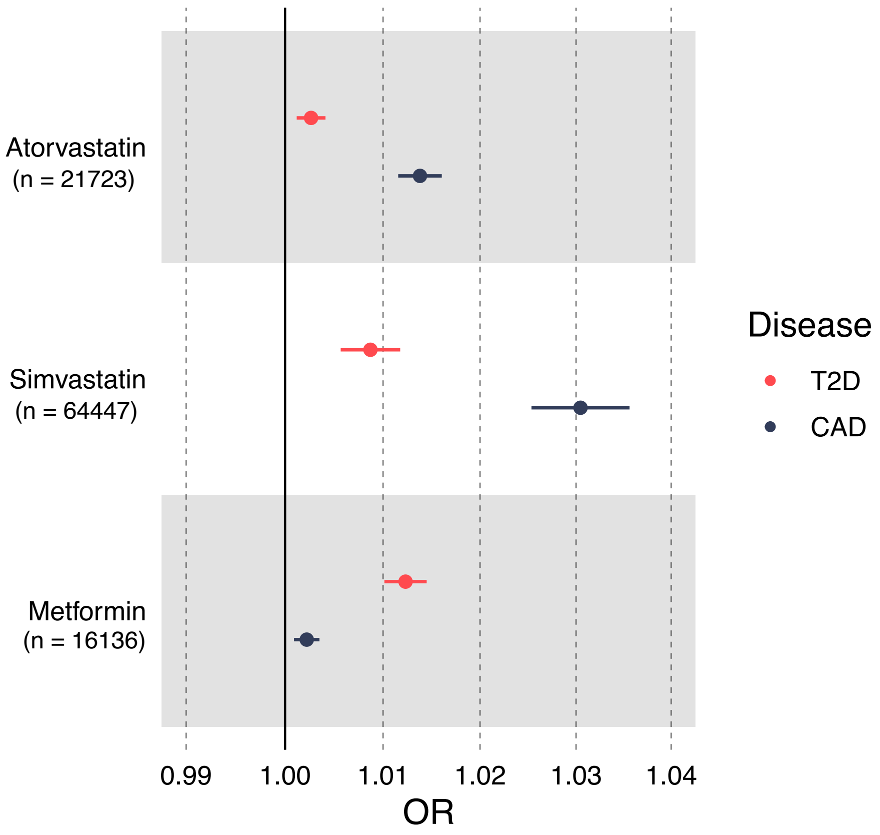


**Supplementary Figure 1:** Effect of T2D and CAD liability on statin and metformin use. Effect estimates are odds ratios when liability to disease is doubled, based on IVW models.

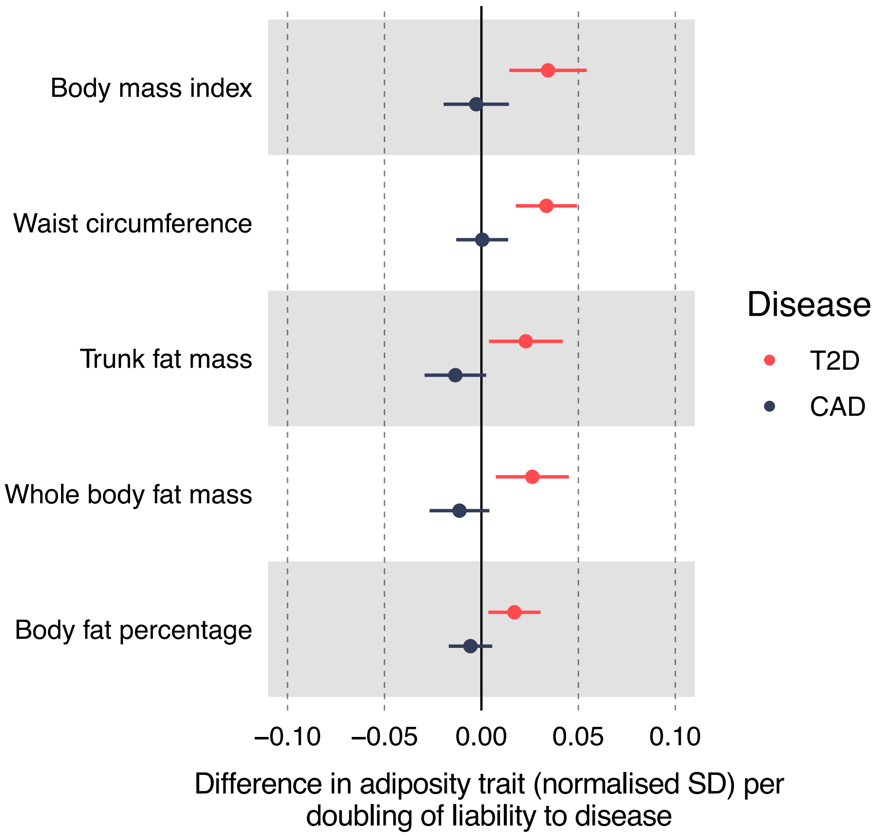


**Supplementary Figure 2:** Effect of T2D and CAD liability on adiposity. Effect estimates are normalised SD unit differences in adiposity trait per doubling of liability to T2D or CAD, based on IVW models.

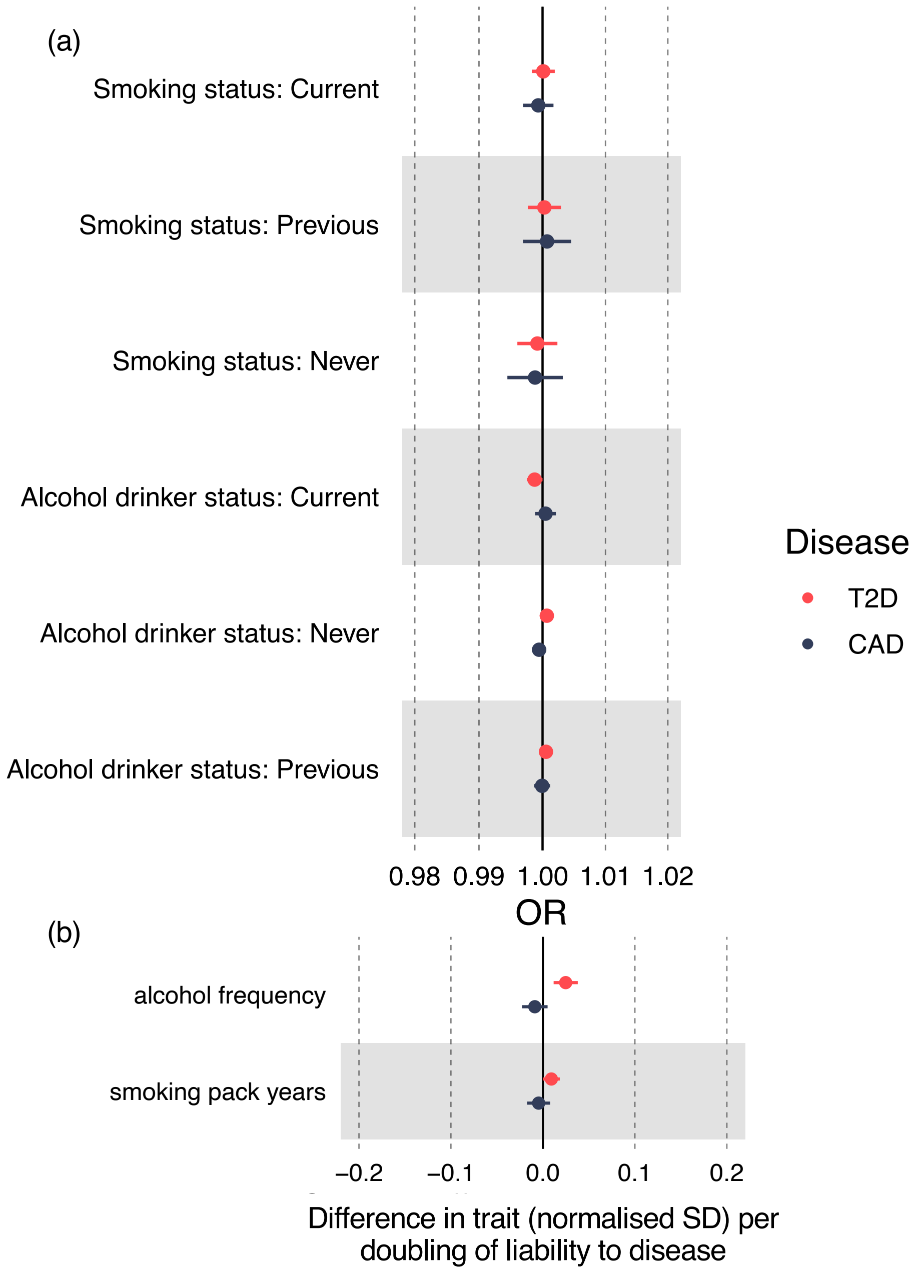


**Supplementary Figure 3**. Effect of T2D and CAD on smoking and alcohol behaviours. Effect estimates are odds ratios when disease liability is doubled in (a) and SD-unit differences in metabolite per doubling of liability to T2D or CAD in (b), based on IVW models.

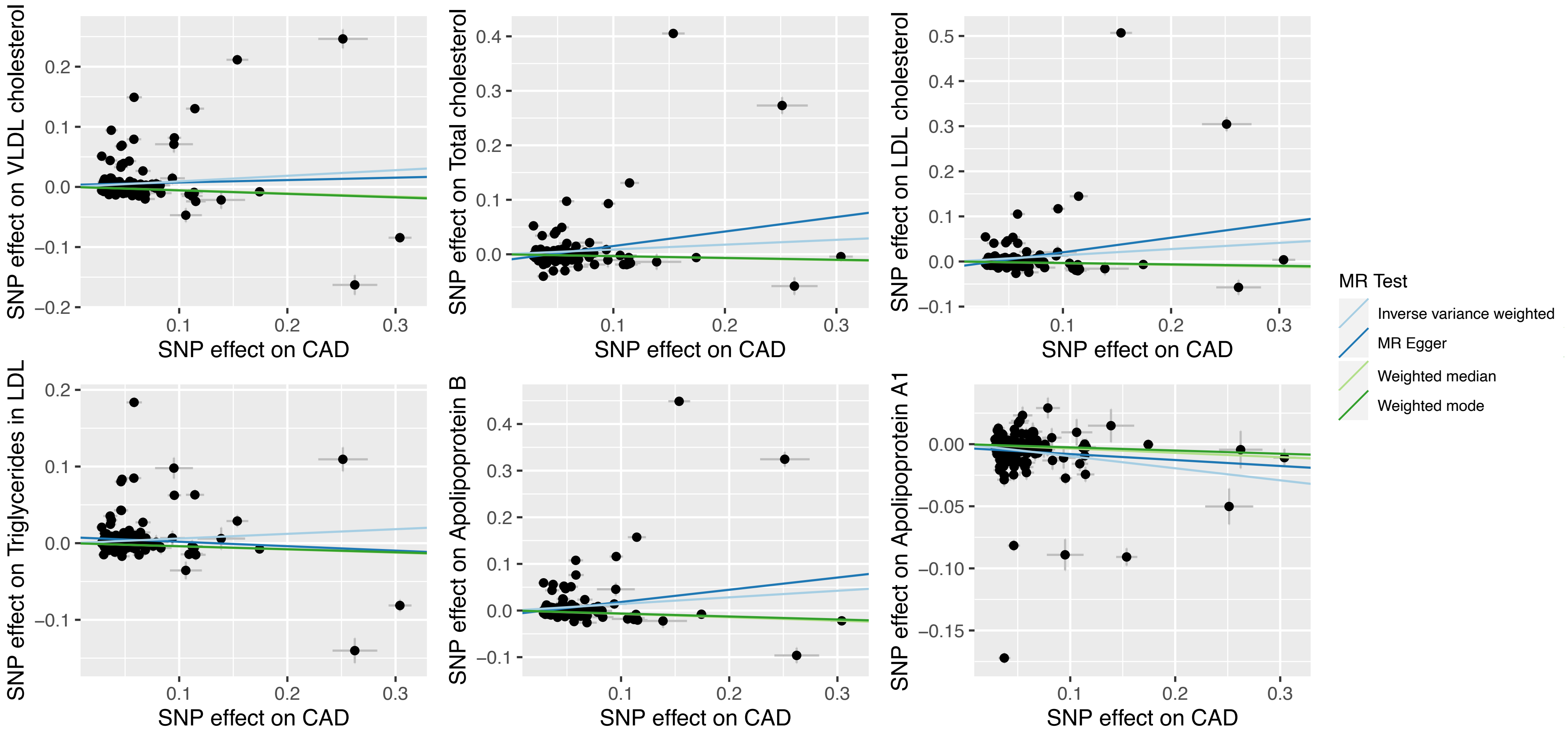


**Supplementary Figure 4**. Scatter plots of individual SNP effects on metabolic traits and CAD.

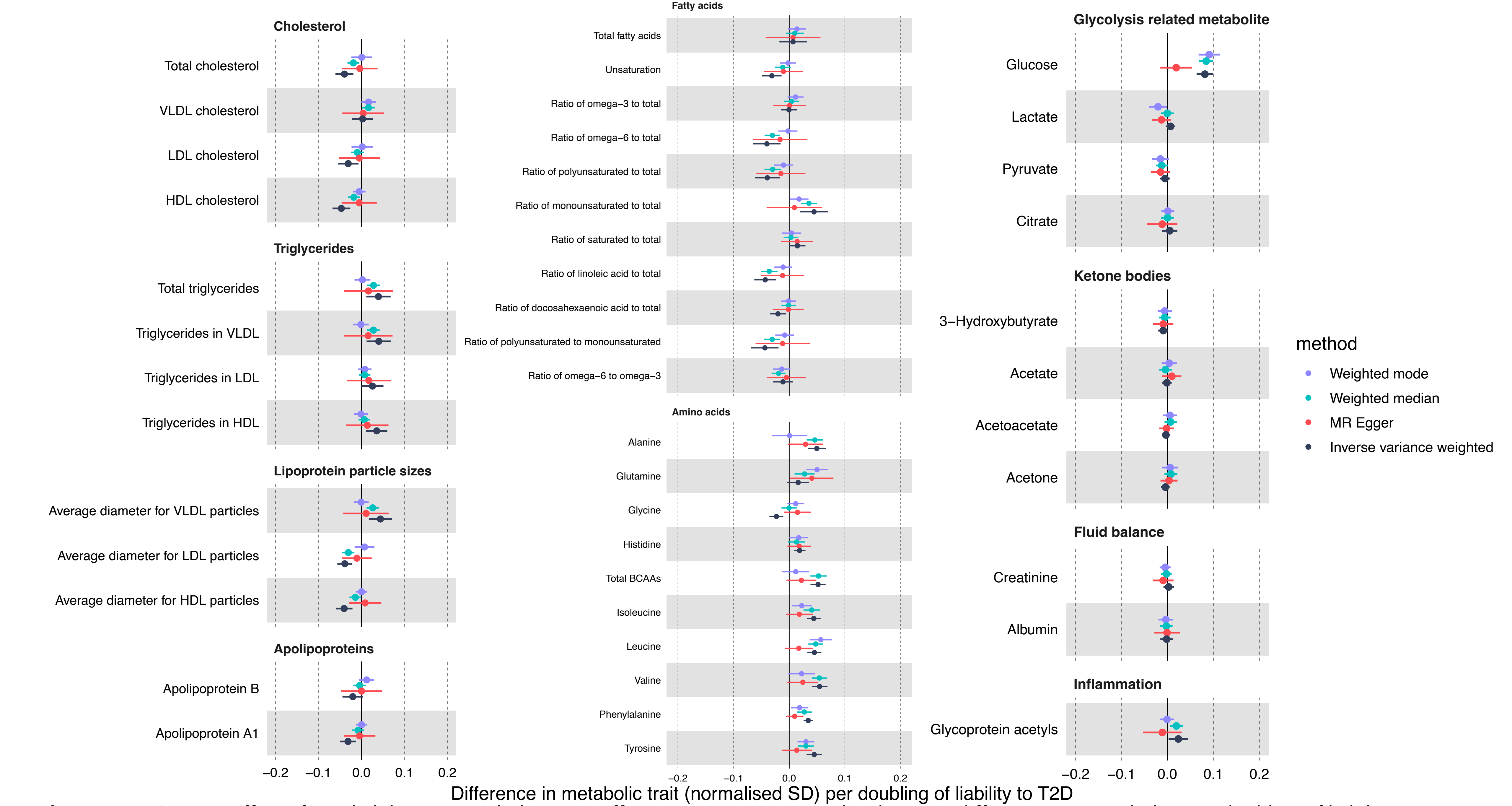


**Supplementary Figure 5:** Effect of T2D liability on metabolic traits. Effect estimates are normalised SD unit differences in metabolite per doubling of liability to T2D based on IVW, MR Egger, weighted median and weighted mode MR models.

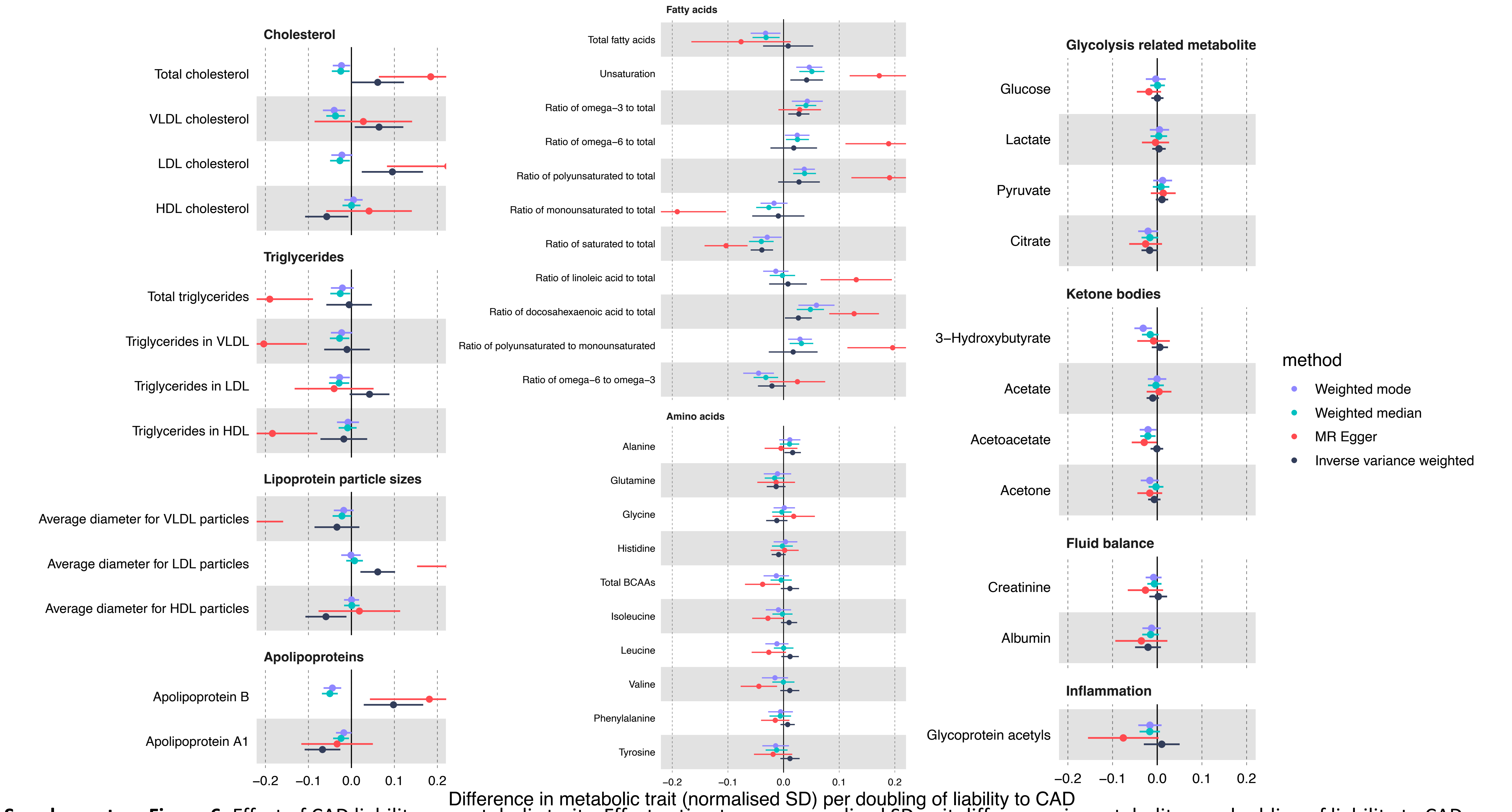


**Supplementary Figure 6:** Effect of CAD liability on metabolic traits. Effect estimates are normalised SD unit differences in metabolite per doubling of liability to CAD based on IVW, MR Egger, weighted median and weighted mode MR models.

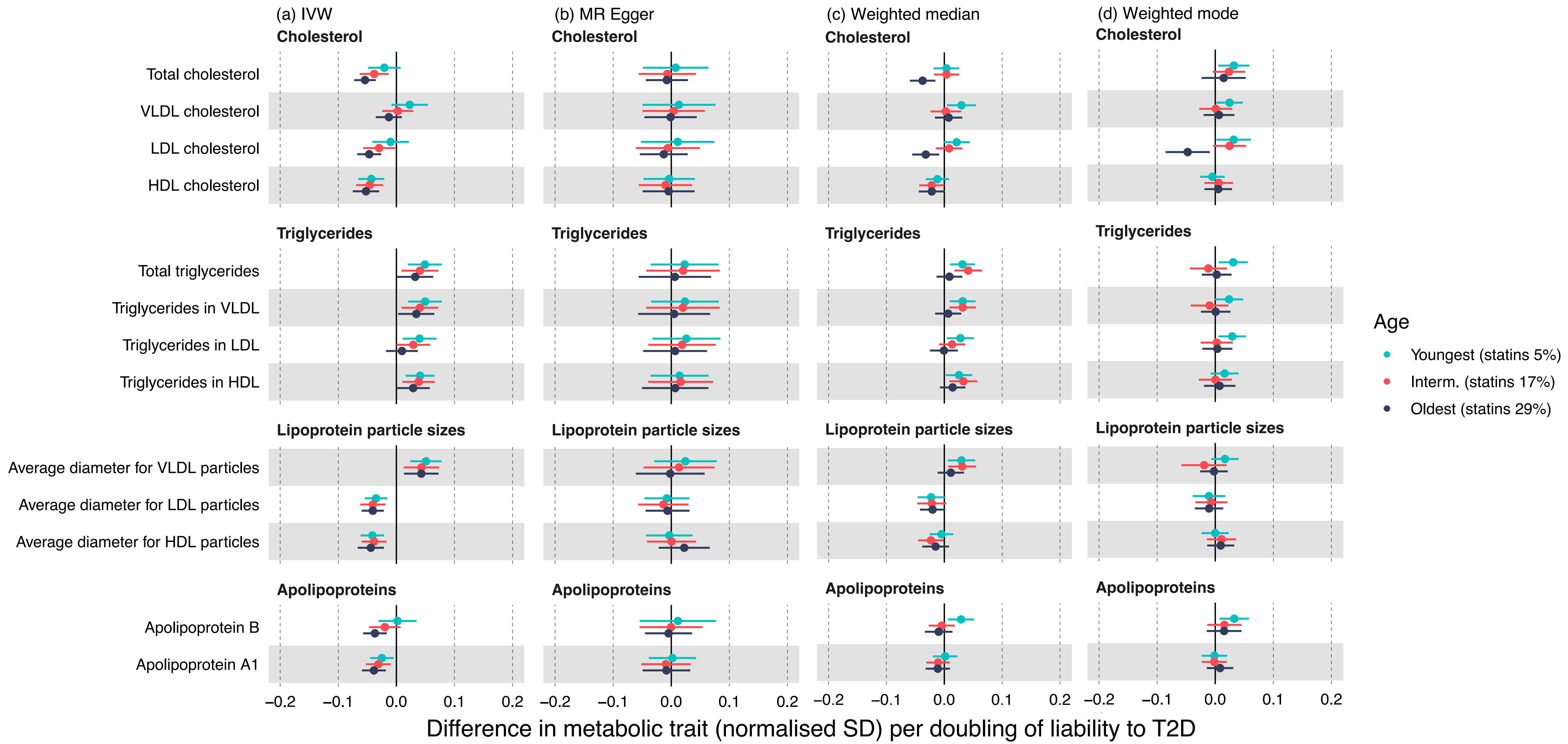


**Supplementary Figure 7:** Effect of T2D liability on lipids and lipoproteins in age tertiles. Effect estimates are normalised SD unit differences in metabolite per doubling of liability to T2D based on (a) IVW, (b) MR Egger, (c) weighted median and (d) weighted mode MR models.

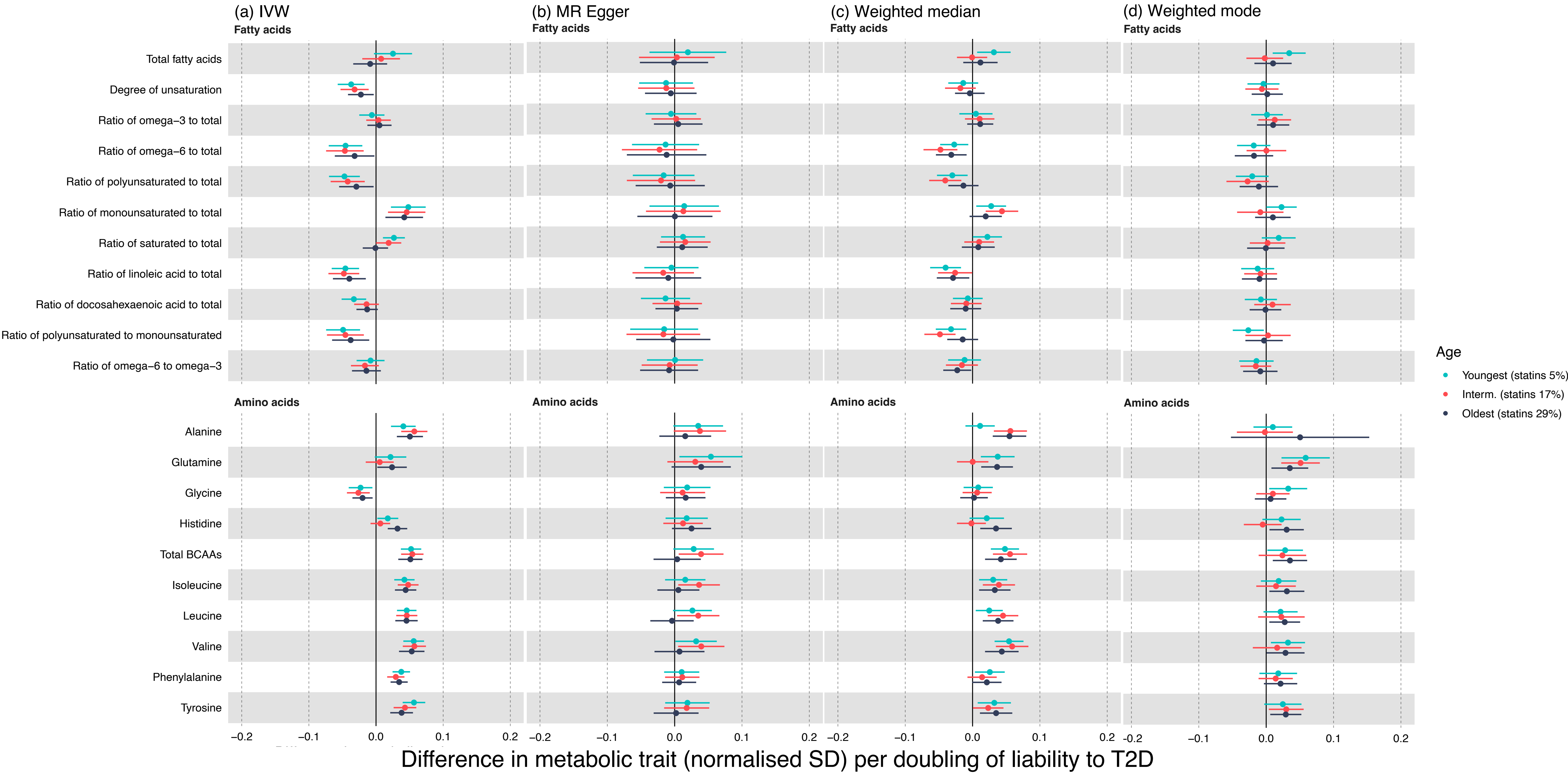


**Supplementary Figure 8:** Effect of T2D liability on fatty acids and amino acids in age tertiles. Effect estimates are normalised SD unit differences in metabolite per doubling of liability to T2D based on (a) IVW, (b) MR Egger, (c) weighted median and (d) weighted mode MR models.

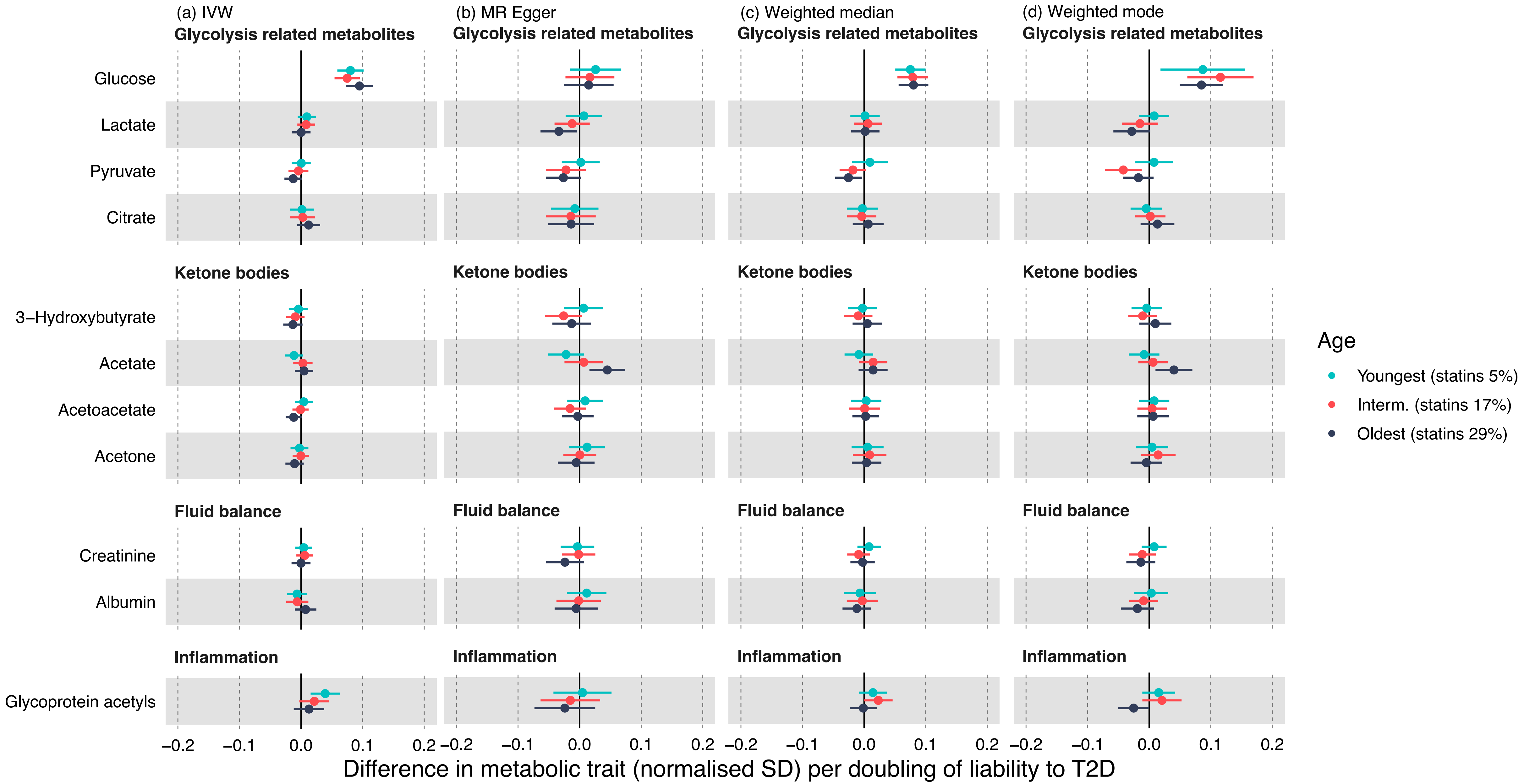


**Supplementary Figure 9:** Effect of T2D liability on glycolysis related metabolites, ketone bodies, fluid balance metabolites and glycoprotein acetyls in age tertiles. Effect estimates are normalised SD unit differences in metabolite per doubling of liability to T2D based on (a) IVW, (b) MR Egger, (c) weighted median and (d) weighted mode MR models.

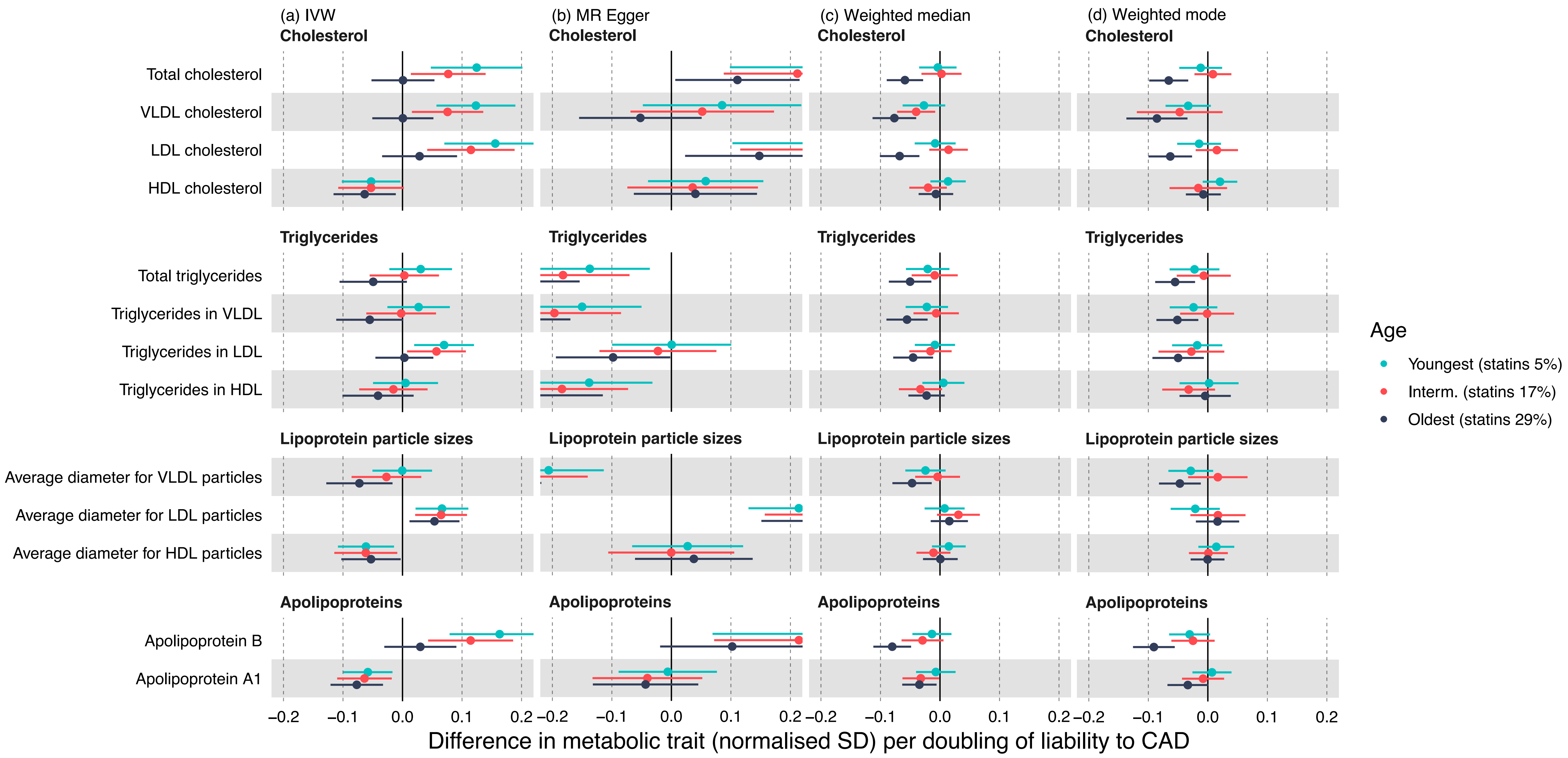


**Supplementary Figure 10:** Effect of CAD liability on lipids and lipoproteins in age tertiles. Effect estimates are normalised SD unit differences in metabolite per doubling of liability to CAD based on (a) IVW, (b) MR Egger, (c) weighted median and (d) weighted mode MR models.

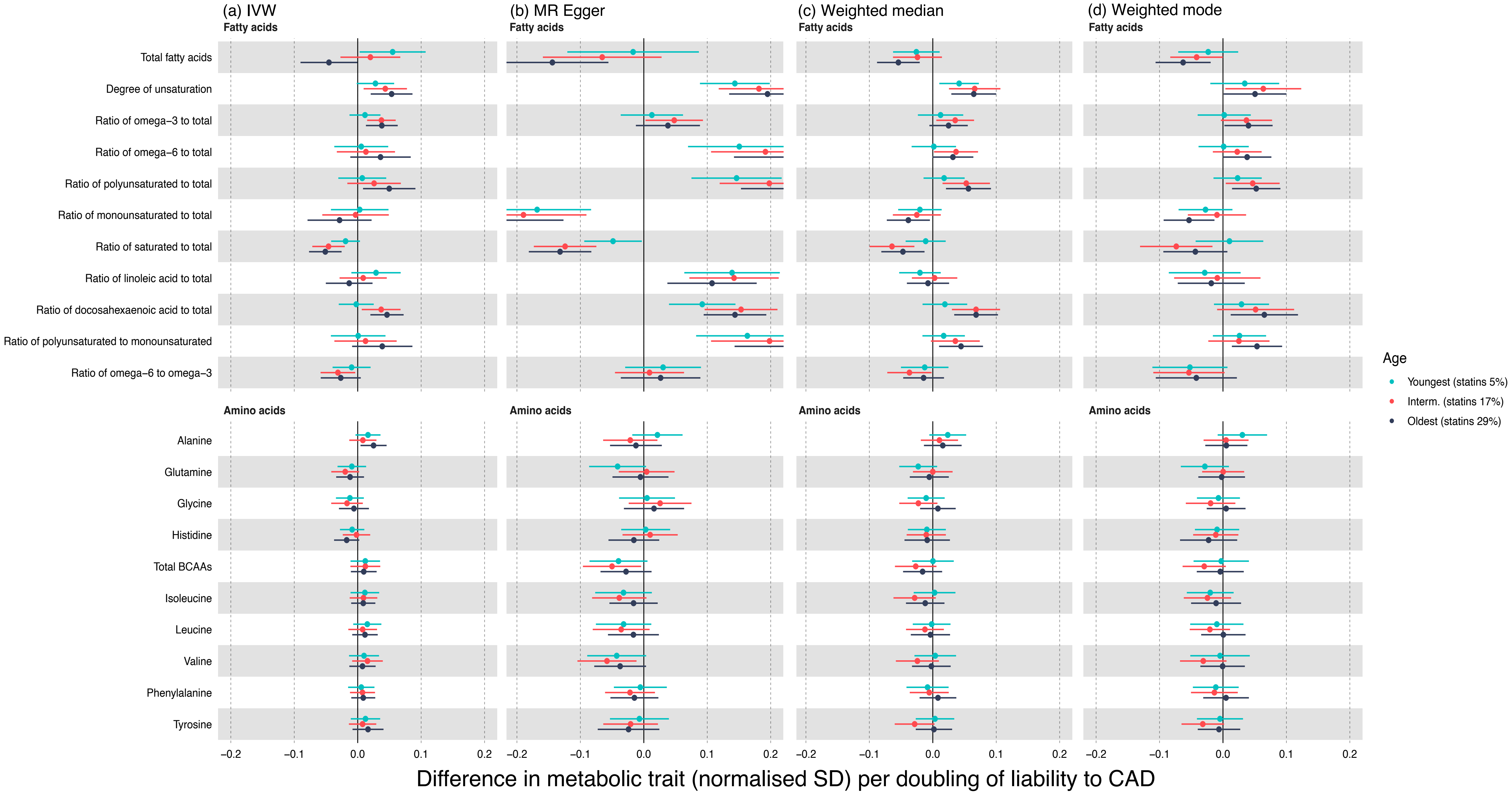


**Supplementary Figure 11:** Effect of CAD liability on fatty acids and amino acids in age tertiles. Effect estimates are normalised SD unit differences in metabolite per doubling of liability to CAD based on (a) IVW, (b) MR Egger, (c) weighted median and (d) weighted mode MR models.

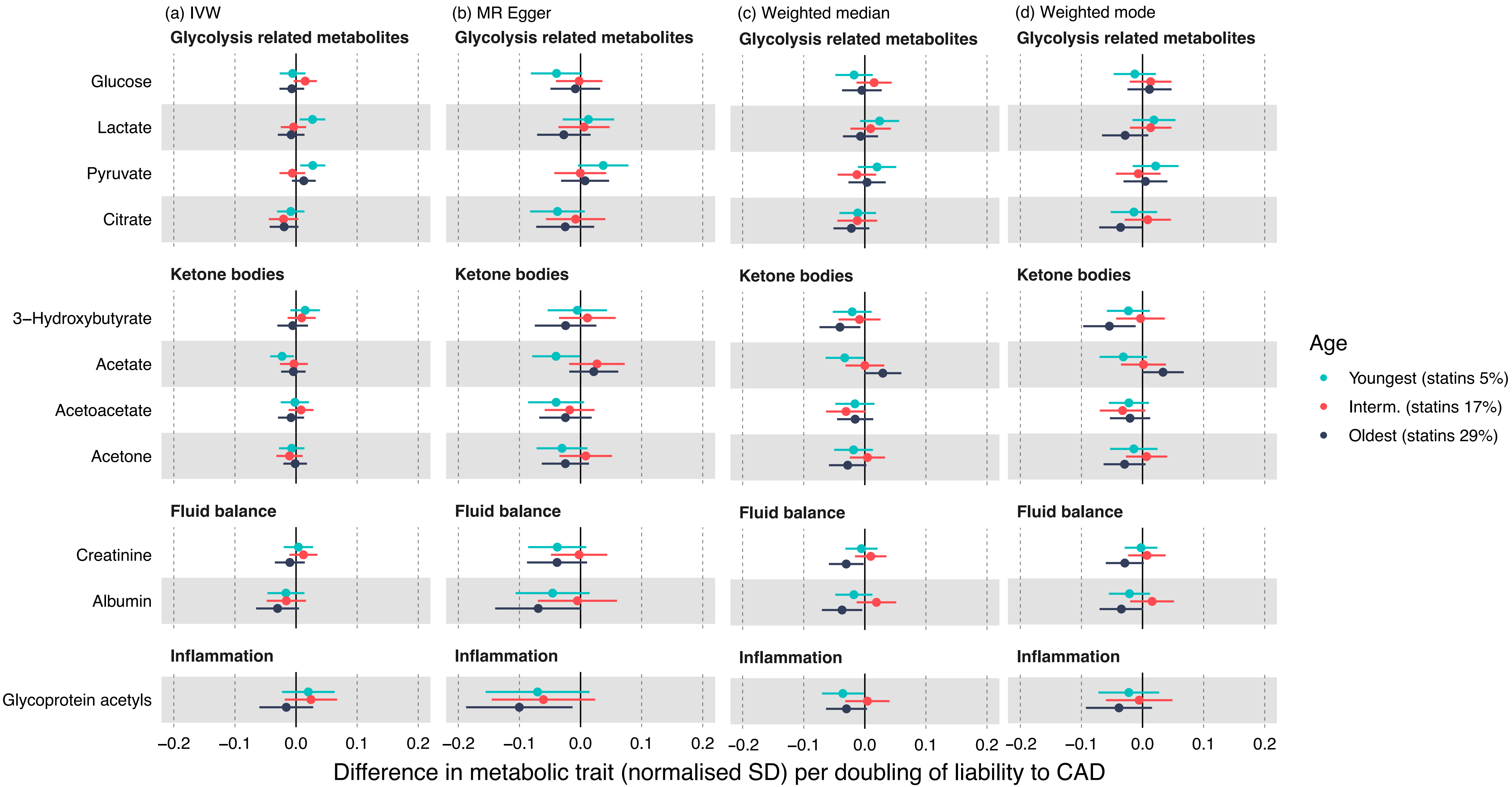


**Supplementary Figure 12:** Effect of CAD liability on glycolysis related metabolites, ketone bodies, fluid balance metabolites and glycoprotein acetyls in age tertiles. Effect estimates are normalised SD unit differences in metabolite per doubling of liability to CAD based on (a) IVW, (b) MR Egger, (c) weighted median and (d) weighted mode MR models.
