## Supplementary File for "Distinct metabolic features of genetic liability to type 2 diabetes and coronary artery disease: a reverse Mendelian randomization study"

**METHODS**

**Additional outcome data on adiposity, smoking, and alcohol consumption**

Summary statistics from GWAS of BMI, waist circumference, trunk fat mass, whole body fat mass, body fat percentage, smoking status, alcohol drinker status, smoking pack-years and alcohol frequency, conducted among European participants of the UK Biobank study were used. Genetic association data were previously generated for these traits using the MRC IEU UK Biobank GWAS pipeline[1], adjusting for age at baseline, sex, and genotyping array.

Standing height was measured without shoes using a Seca 202 device and weight was measured with heavy outer clothing removed, using the Tanita BC418MA body composition analyser (bioelectrical impedance). This same device was used to estimate trunk fat mass in kilograms, whole body fat mass in kilograms, and body fat percentage. Body mass index (BMI) was calculated from dividing weight in kilograms by the square of height in metres. Waist circumference (WC) was measured in centimetres at the umbilicus using a non-stretchable Wessex tape measure.

Alcohol drinking status (never, previous or current) was measured (N=336,965) via a questionnaire. Alcohol intake frequency was measured (N=462,346) via a questionnaire where they were asked “About how often do you drink alcohol?” and answered from the following options: “Daily or almost daily”, “Three or four times a week”, “Once or twice a week”, “One to three times a month”, “Special occasions only”, “Never” or “Prefer not to answer”. Smoking status (never, previous, current) was also measured via questionnaire (N=336,024). Smoking pack years was calculated for 142,387 participants that have ever smoked according to their smoking status response. The general definition of a pack year is the number of cigarettes smoked per day, divided by twenty, multiplied by the number of years of smoking. Number of years of smoking was calculated by subtracting the age the participant started smoking from the age they stopped.

**Proxy SNPs**

Where possible, when exposure SNPs were not available in the outcome GWAS, proxy SNPs were identified using 1000 genomes sample data as SNPs in LD (within 10,000kb, r^2^=0.8) with the requested SNP. The number of proxy SNPs recruited differed for each outcome (maximum number of proxies for any one outcome = 8).

**RESULTS**

**Instrument count and strength**

167 independent genetic variants were associated with T2D at P < 5 x 10^-8^, 159 of which were present in the outcome GWAS summary statistics and were used in MR analyses. For the medication use and other outcomes, between 155 and 159 SNPs were available for each and used for MR analyses. One hundred and forty-five genetic variants were associated with CAD at P < 5 x 10^-8^, 140 of which were available for each metabolic trait and thus used to instrument liability to CAD. For the medication use and other outcomes, 136-141 SNPs were available for each. The F statistics for the genetic instruments for T2D and CAD were 66.8 and 59.7, respectively, indicating instrument strength above the recommended minimum level of 10 to avoid weak instrument bias.[2]
